# Immunogenicity and Safety of a SARS-CoV-2 Inactivated Vaccine in Healthy Adults Aged 18-59 years: Report of the Randomized, Double-blind, and Placebo-controlled Phase 2 Clinical Trial

**DOI:** 10.1101/2020.07.31.20161216

**Authors:** Yanjun Zhang, Gang Zeng, Hongxing Pan, Changgui Li, Biao Kan, Yaling Hu, Haiyan Mao, Qianqian Xin, Kai Chu, Weixiao Han, Zhen Chen, Rong Tang, Weidong Yin, Xin Chen, Xuejie Gong, Chuan Qin, Yuansheng Hu, Xiaoyong Liu, Guoliang Cui, Congbing Jiang, Hengming Zhang, Jingxin Li, Minnan Yang, Xiaojuan Lian, Yan Song, Jinxing Lu, Xiangxi Wang, Miao Xu, Qiang Gao, Fengcai Zhu

**Author notes:** Footnote The first six authors, YZ, GZ, HP, CL, BK, and YH, contribute equally to the manuscript and are listed as the first authors. The last five authors, JL, XW, MX, QG, and FZ, contribute equally to the correspondence and are listed as the corresponding authors.

## Abstract

**BACKGROUND:** The top priority for the control of COVID-19 pandemic currently is the development of a vaccine. A phase 2 trial conducted to further evaluate the immunogenicity and safety of a SARS-CoV-2 inactivated vaccine (CoronaVac).

**METHODS:** We conducted a randomized, double-blind, placebo-controlled trial to evaluate the optimal dose, immunogenicity and safety of the CoronaVac. A total of 600 healthy adults aged 18-59 years were randomly assigned to receive 2 injections of the trial vaccine at a dose of 3 μg/0.5 mL or 6 μg /0.5mL, or placebo on Day 0,14 schedule or Day 0,28 schedule. For safety evaluation, solicited and unsolicited adverse events were collected after each vaccination within 7 days and 28 days, respectively. Blood samples were taken for antibody assay.

**RESULTS:** CoronaVac was well tolerated, and no dose-related safety concerns were observed. Most of the adverse reactions fell in the solicited category and were mild in severity. Pain at injection site was the most frequently reported symptoms. No Grade 3 adverse reaction or vaccine related SAEs were reported. CoronaVac showed good immunogenicity with the lower 3 μg dose eliciting 92.4% seroconversion under Day 0,14 schedule and 97.4% under Day 0,28 schedule. 28 days after two-dose vaccination, the Nab levels of individual schedules range from 23.8 to 65.4 among different dosage and vaccination schedules.

**CONCLUSIONS:** Favorable safety and immunogenicity of CoronaVac was demonstrated on both schedules and both dosages, which support the conduction of phase 3 trial with optimum schedule/dosage per different scenarios.

## BACKGROUND

In January 2020, outbreaks of coronavirus disease in 2019 (COVID-19) caused by severe acute respiratory syndrome coronavirus 2 (SARS-CoV-2) escalated rapidly, and since then COVID-19 cases have been reported in over 200 countries and territories. The pandemic continues to spread unabated affecting the health and changing the lifestyles of people globally.^1^ To reduce the disease burden and stop the community-wide transmission of COVID-19 across the globe, specific therapeutic agents or vaccines are urgently needed. Till now, more than 120 vaccine candidates have been reported to be under development and at least 23 have progressed to the clinical evaluation stage.^2^

The inactivated SARS-CoV-2 vaccine with aluminum hydroxide developed by Sinovac Life Sciences Co.. Ltd., also known as CoronaVac, has been shown to be safe and could induce SARS-CoV-2 specific neutralizing antibodies in mice, rats, and nonhuman primates.^3^ On the basis of the results obtained from our phase 1 trial, no safety concerns have been identified. Notably, immunization of CoronaVac induced immune responses against SARS-CoV-2 in adults. Here, we report the results of the phase 2 trial.

## METHODS

### TRIAL DESIGN AND OVERSIGHT

This double-blind, randomized and placebo-controlled phase 2 clinical trial based on a seamless design was registered at clinicaltrials.gov (NCT04352608) and was conducted in Suining County. Jiangsu Province. China. Detailed information about the trial has been provided in our previous phase 1 study. The trial protocol and the informed-consent form were approved by the ethics committee of the Jiangsu Provincial Center for Disease Control and Prevention (JSCDC). This clinical trial was conducted in accordance with the Chinese regulatory requirements and the standards of good clinical practice.

Before enrollment, written informed consent was obtained from each participant. The main exclusion criteria included high-risk epidemiological history, positive IgG, IgM or nucleic acid test of pharyngeal or anal swab, axillary temperature >37.0, allergy to a vaccine component, and other unsuitable conditions.

A total of 600 healthy adults aged 18-59 years were randomly assigned into 3 groups in a ratio of 2:2:1 to receive 2 injections of the trial vaccine at a dose of 3 μg/0.5 mL or 6 μg /0.5mL, or placebo on a Day 0,14 schedule or a Day 0,28 schedule, according to a random list generated by an independent statistician..

### VACCINE

The vaccine candidate was an inactivated SARS-CoV-2 whole virion vaccine with aluminium hydroxide as adjuvant (CoronaVac) developed by Sinovac Life Sciences Co., Ltd. SARS-CoV-2 virus was propagated in Vero cells and harvested. The harvested virus was inactivated using β-propiolactone and further purified. The bulk vaccine material obtained from this step was then adsorbed onto aluminium hydroxide and formulated with phosphate-buffered saline (PBS) and sodium chloride as inactivated final product. The dosage of 3 μg/0.5 mL and 6 μg/0.5mL were adopted in this study. Whereas the placebo contained aluminum hydroxide diluents with no antigen. Both were administered intramuscularly on the schedule of Day 0,14 or Day 0,28.

### SAFETY ASSESSMENT

For safety evaluation of CoronaVac, the participants who received at least one dose of vaccination was included. All vaccinated subjects were observed for immediate adverse events (AEs) on-site for at least 30 minutes after each administration. Diary cards were issued to the participants to record the solicited AEs (e.g, pain, induration, swelling, redness, rash, pruritus) occurring on day 0~7 and unsolicited AEs (e.g, fever, acute allergic reaction, skin and mucosa abnormality, diarrhea, anorexia, vomiting, nausea, muscle pain, headache, cough, fatigue) occurring on day 0~28. Data on serious adverse events (SAEs) were collected throughout the trial. All AEs were assessed for severity, and the relationship to vaccination was decided by investigators before unblinding.

### IMMUNOGENICITY

To assess immune response, blood samples were collected from each participant different time points (0/28/42^th^ day for Day 0,14 schedule, and 0756^th^ day for Day 0,28 schedule). The ability of the antibodies present in the blood sample to bind the receptor binding domain (RBD) of SARS-CoV-2 was assessed by enzyme-linked immunosorbent assay (ELISA). A dilution of 1:160 was considered as a positive cutoff value. We also measured neutralizing antibody titer (Nab) using a modified cytopathogenic effect assay. A titer of 1:8 or higher indicated seropositivity. Seroconversion was defined as a change from seronegative (<1:8) to seropositive (≥1:8) or a 4-fold increase from baseline titers if seropositive.

The neutralizing antibody assay was performed by Chinese National Institutes for Food and Drug Control, and the ELISA was performed by Sinovac Biotech.

### NEGATIVE STAIN

Virus particles of vaccine used for phase 1 and 2 were diluted to a concentration of 0.04 mg/mL, deposited on a glow-discharged carbon-coated copper grid (Electron Microscopy Sciences) and after 1 min, washed twice with buffer (20 mM Tris, 200 mM NaCl, pH 8.0), and stained with 1% phosphotungstic acid (pH 7.0) for 1 min. Then the grid was imaged at room temperature using FEI Tecnai Spirit electron microscope (Thermo Fisher Scientific) operated at an acceleration voltage of 120 kV.

### STATISITICAL ANALYSIS

Safety evaluation was performed on participants who received at least 1 dose of the vaccine or placebo by comparing the overall incidence rate of solicited and unsolicited AEs among relevant groups. Immunogenicity assessment was performed on the per-protocol set (PPS). The seroconversion rate was defined as a change from seronegative to seropositive or a 4-fold increase from baseline titers if seropositive. The titer distributions were described with reverse cumulative distribution curves and were tested with the nonparametric Kruskal-Wallis test over the groups.

The Pearson Chi-square test or Fisher’s exact test was adopted for the analysis of binary outcomes. Clopper-Pearson method was used to compute the 95% confidence intervals (CIs) of the binary outcome. ANOVA method was utilized to compare the GMTs among groups. Hypothesis testing was two-sided with an alpha value of 0.05. Analyses were conducted by SAS 9.4 (SAS Institute, Cary, NC, USA).

## RESULTS

### STUDY POPULATION

From 29 April to 5 May 2020, 600 subjects were enrolled and randomly assigned to receive first of the CoronaVac or placebo dose. All subjects were included into the safety assessment. During this trial, 297 subjects put on Day 0,14 schedule and 294 subjects following Day 0,28 schedule were included in the per-protocol cohort for immunogenicity analysis. These subjects received the 2 injections, attended all visits and gave planned blood sample. Information about study enrollment, randomization, and vaccination is shown in Fig. S1.

Baseline demographic characteristics at enrollment were similar among these groups in terms of sex, mean age, height, and weight (Table 1).

**Table 1.** Baseline Characteristics of the Study Participants.*

| Characteristics | 3 µg Group | 6 µg Group | Placebo | *P* |
| --- | --- | --- | --- | --- |
| Day 0,14 schedule |  |  |  |  |
| N | 120 | 120 | 60 |  |
| Age (years) | 42.0±10.2 | 42.4±9.0 | 43.6±7.6 | 0.5543 |
| Gender (male/female) | 54/66 | 48/72 | 25/35 | 0.7305 |
| Height (m) | 1.7±0.1 | 1.6±0.1 | 1.6±0.1 | 0.3864 |
| Body weight (kg) | 67.8±11.7 | 68.7±11.5 | 68.4±10.9 | 0.8258 |
| BMI (kg/m<sup>2</sup>) | 24.9±3.6 | 25.5±3.2 | 25.5±3.0 | 0.2930 |
| Day 0,28 schedule |  |  |  |  |
| N | 120 | 120 | 60 |  |
| Age (years) | 41.5±9.6 | 40.6±9.9 | 44.3±8.4 | 0.0472 |
| Gender (male/female) | 63/57 | 63/57 | 30/30 | 0.9417 |
| Height (m) | 1.7±0.1 | 1.7±0.1 | 1.7±0.1 | 0.9433 |
| Body weight (kg) | 70.0±11.8 | 70.0±12.2 | 72.1±12.2 | 0.4704 |
| BMI (kg/m2) § | 25.2±3.1 | 25.2±3.3 | 26.1±3.1 | 0.1741 |

\* Plus-minus values are means ±SD.
§ BMI=body mass index.

### ADVERSE REACTIONS

For subjects in Day 0,14 schedule, the incidence rates of adverse reactions in 6 μg, 3 μg and placebo group were 35.0%, 33.3% and 21.7%, respectively; while the corresponding incidence rates were 19.2%, 19.2% and 18.3% in Day 0,28 schedule, respectively. Within each schedule, there was no significant difference in the occurrence of adverse reactions among all vaccine and placebo groups (Fig. 1). Most of the adverse reactions were solicited adverse reactions and mild in severity. After each injection, pain at the injection site was the most frequently reported local symptoms, which reported in 61 subjects (20.3%) on Day 0,14 schedule and 31 subjects (10.3%) on Day 0, 28 schedule. (Additional detailed results related to adverse reactions are available in Table S1).

We did not observe any Grade 3 adverse reaction. Most reported adverse reactions resolved within 72 hours after vaccine administration. During the follow-up period, 3 SAEs were reported from 3 subjects and neither was vaccine related.

**Figure 1.**
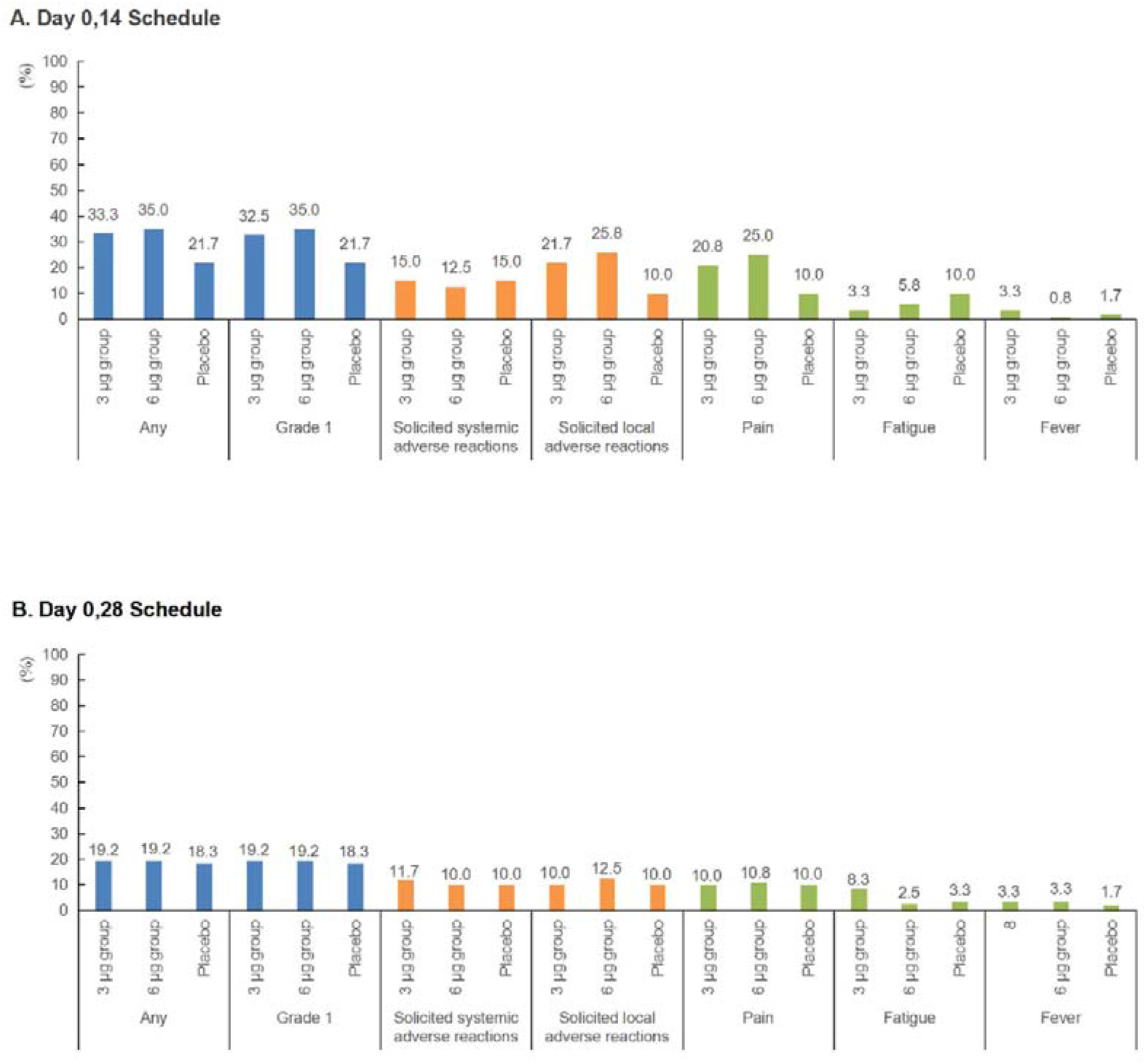
Incidence rates of adverse reactions among different groups in phase 2. (A) The incidence rates of adverse reactions among different groups with a Day 0,14 schedule. (B) The incidence rates of adverse reactions among different groups with a Day 0,28 schedule.

### IMMUNOGENICITY

At baseline, all the 600 subjects were seronegative (with Nab titers of <1:8); but the seroconversion rates increased over 90% during the later stages of the trial. Within each dosage, there was no significant difference in the seroconversion rates between Day 0,14 and Day 0,28 schedule. For the antibody response against the receptor binding domain, similar results were observed (Table S2). No changes in seropositivity frequencies and GMTs from baseline were found for the placebo group.

For subjects on Day 0,14 schedule, the GMT increased to 34.5 (95% CI, 28.5 to 41.8) and 27.6 (95% CI, 22.7 to 33.5) in 6 μg and 3 μg group, respectively, and remained stable after 28 days from the second injection (Fig. 2A). The neutralizing antibody titers for subjects on Day 0, 28 schedule increased significantly 28 days after the second injection, when compared to those of subjects on Day 0,14 schedule within each dosage group. Almost similar trends like those observed for the neutralizing antibody were observed during the evaluation of the IgG antibody level (Fig. 2B). In addition, the neutralizing antibody titers significantly decreased with increasing age (Fig. 2C and 2D); younger subjects tended to have a higher level of neutralizing antibody titers.

**Figure 2.**
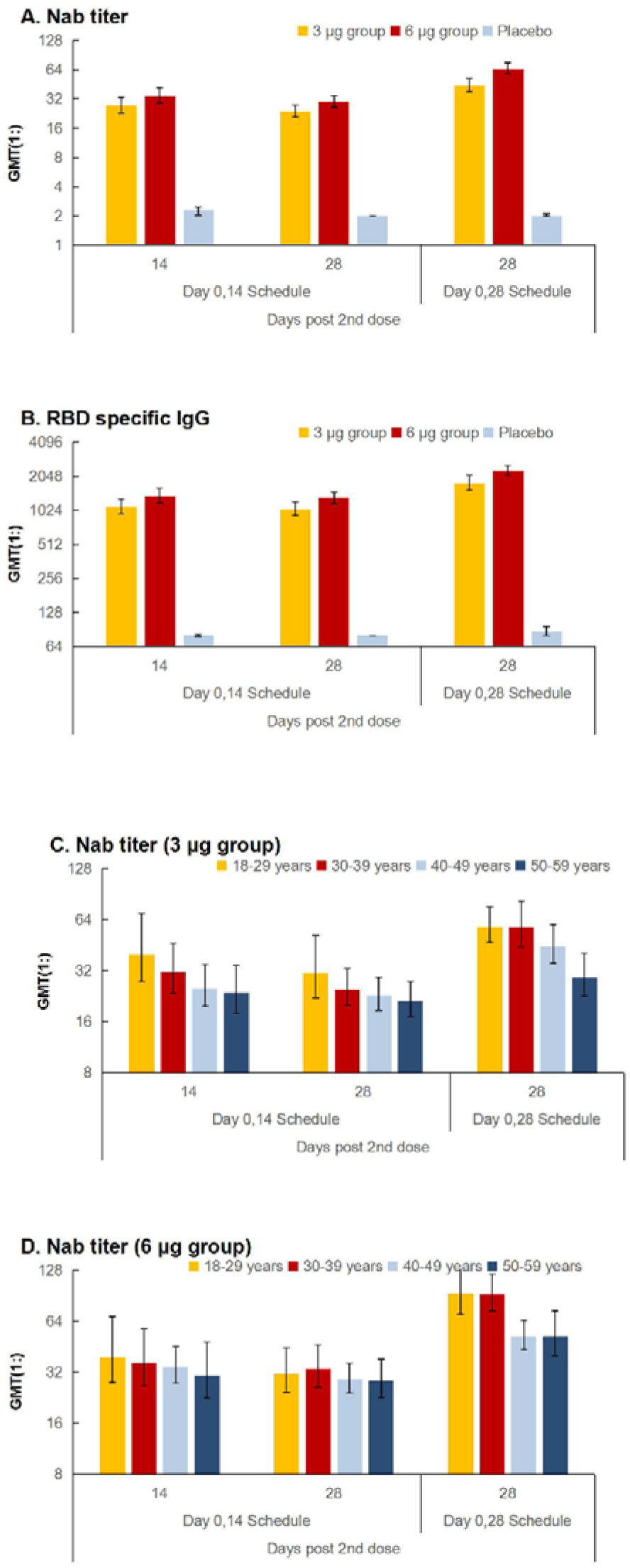
Antibody Response in the Per-Protocol Cohort. (A) The neutralizing antibody titer in all participants 14 and 28 days after second dose in Day 0,14 schedule and 28 days after second dose in Day 0,28 schedule. (B) The RBD specific IgG antibody titer in all participants 14 and 28 days after second dose in Day 0,14 schedule and 28 days after second dose in Day 0,28 schedule. (C) The neutralizing antibody titer among different age-groups at different time points from all participants that received 3 μg vaccine. (D) The neutralizing antibody titer among different age-group at different time points from all participants that received 6 μg vaccine.

**Figure 3.**
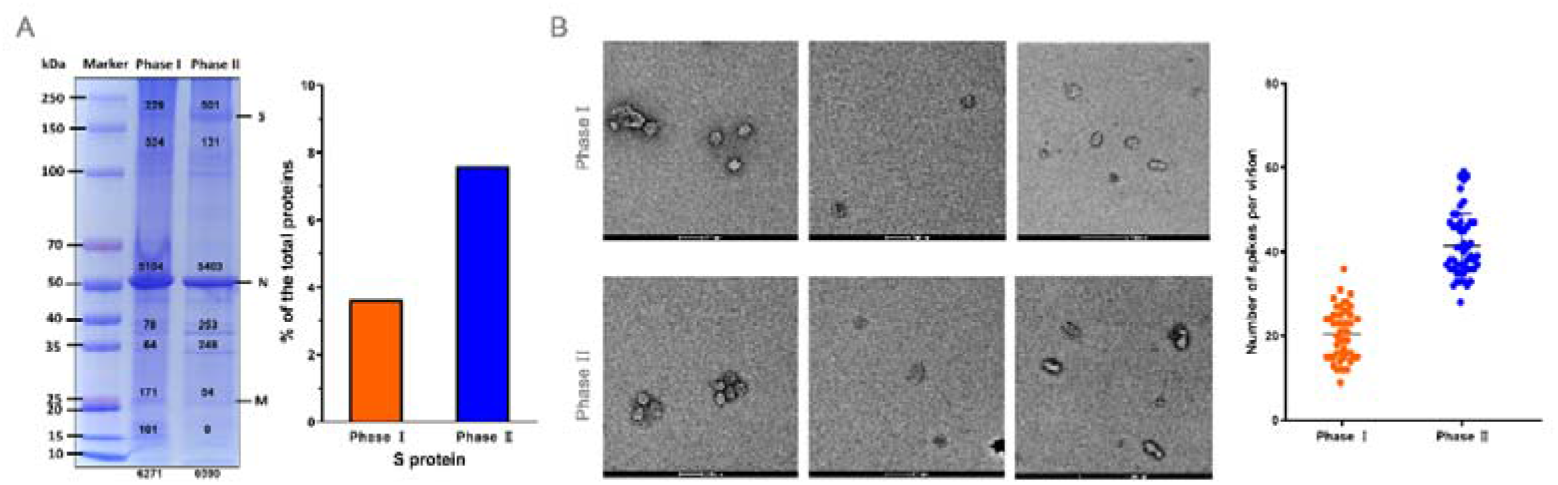
The proportion of Spikes in CoronaVac used for phase 1 and 2 vaccine evaluation. (A) Protein composition analysis of CoronaVac samples from phase I and II by a NuPAGE 4-12% Bis-Tris gel, followed by whole-gel protein staining using Coomassie Blue gel staining reagent (45% methanol, 10% glacial acetic acid, 0.25% Coomassie Blue R-250). The viral protein bands of vaccine strain used for phase I and II were quantified by densitometry using ImageJ software with values depicted in the gel. The proportions of spikes to the total proteins in each gel lane in CoronaVac samples used forof phase 1 and 2 were calculated separately. (B) Representative negative staining images of the CoronaVac samples used in phase 1 and 2 trials. Three images were randomly selected for each phase. Grouped scatter plot showing the numbers of Spikes on two-dimensional projections of randomly selected 50 virions of CoronaVac samples used for phase I (left) and phase II (right), respectively.

## DISCUSSION

This trial demonstrated that the 2 doses of different dosage of CoronaVac were well tolerated and immunogenic in healthy adults aged 18-59 years. The incidence rates of adverse reactions in the 6 μg and 3 μg group were comparable, indicating that there was no dose-related aggravating concern on safety. Furthermore, no SAEs related to vaccine occurred, and most adverse reactions reported were generally assessed to be mild. The safety profile of CoronaVac is comparable to that observed in our phase 1 clinical trial [see the coordinated submission], and to other inactivated vaccine formulations manufactured by Sinovac.^4,5^ Compared with other COVID-19 vaccine candidates, the incidence rate of fever was relatively low in our clinical trial, which further indicates that CoronaVac was well tolerated.^6–10^

It’s worth noting that the immune responses elicited in phase 2 were much better than those recorded in phase 1, with seroconversion rates over 90%. Our preclinical investigations had revealed that cell culture technology closely correlated with viral propagation and affected viral morphology, protein composition and prefusion conformation of spikes.^3^ In both preclinical study and phase 1 trials, a 50-liter culture of Vero cells grown in the Cell Factory system was used, while an optimized process for growing cells using a highly automated bioreactor, where cell culture parameters like dissolved oxygen, pH, and CO_2_/O_2_ gas levels, were controlled precisely, was developed for producing the CoronaVac for phase 2 trial. To deduce the reasons underlying the enhanced protective immune responses observed in phase 2 trial, we examined the molecular differences between the CoronaVac used in phase 1 and 2 trials. Protein composition analysis of the purified inactivated SARS-CoV-2 virions indicated that the bioreactor-produced CoronaVac possessed higher redundancy of intact spike protein (~180 kDa) when compared to the Cell Factory-yielded CoronaVac (Fig. 3A). Quantitative analysis showed that the intact spike protein accounted for ~7% and ~ 3.7 of total protein mass used in phase 1 and 2 trials, respectively. Electron microscopic examination of the samples further verified that the average number of spikes per virion of the viral sample used in phase 2 trial was almost double to those used in phase 1 trial (Fig. 3B). These observations indicated that CoronaVac used in phase 2 trial contained more *bona fide* immunogens, which explains its better protective immune responses, highlighting the importance of developing an optimum manufacturing process and the integration of multiple-disciplinary techniques, such as genomics and structural biology to support a new era of precision vaccinology.

After two-dose vaccination, immune responses induced by Day 0,28 schedule was above the value of Day 0,14 schedule regardless of the dosage of the vaccine, which was consistent with our anticipation. By using Day 0,14 schedule, antibody response could be induced within a relatively short time period, and this schedule could be introduced to an emergency use and is of vital importance to handle COVID-19 pandemic situation. Regarding the Day 0,28 schedule, robust antibody response is generated and longer persistence could be expected, which supports the need for a routine use under the low incidence rate of COVID-19.

Nabs play an important role in virus clearance and have been considered as a key immune correlate for protection or treatment against viral diseases. Twenty-eight days after the two-dose vaccination, the Nab levels of individual schedules range from 23.8 to 65.4 in phase 2, which was lower than those of convalescent patients tested by the same method in the same laboratory, of which the Nab average level was 163.7.^11^ We assume the antibody level could provide satisfying protection against COVID-19 disease based on three reasons. Firstly, most of the surrogate endpoints based on neutralizing antibodies ranges from 8-24, such as EV71 and Varicella vaccines.^12,13^ Secondly, experience from our preclinical study indicated that the neutralizing antibody titers of 1:24 elicited in macaques models conferred complete protection against SARS-CoV-2. Thirdly, several studies revealed that antibody responses generated from natural infection may decreased significantly, such as SARS-Cov-2, SARS-CoV and MERS-CoV,^14–16^ however, recrudesce of these patients has been rarely reported, which indicated that the immunological memory might play an important role of prevention of re-infections.

Moreover, one prospective goal of our preclinical study and clinical trials was to establish a vaccine-induced surrogate of protection. Compared with vaccine inducing high level antibody, those inducing lower antibody level are more likely to produce evidence on surrogate of protection. Under above assumptions, the dosage of 3 μg with Day 0,14 or Day 0,28 schedule is adopted in our phase 3 trial.

When comparing antibody levels between age-groups, it should be noted that the neutralizing antibody titers significantly decreased with increasing age. These results are consistent with epidemiological trends observed in COVID-19 patients; those with moderate or severe symptoms tend to be elderly.^17^ These results suggest that escalated dosage or extra dose of CoronaVac might be needed in elderly.

Several limitations of this trial should be noted. Firstly, we only assessed the humoral immunity in phase 2 trial, and more evaluation focus on response of Th1 and Th2 is ongoing. Secondly, we only reported immune response data on healthy adults, and do not include data on more susceptible populations, such as elderly or with comorbidity; and also the immune persistence is not available yet, which need to be further studied.

Thirdly, we didnt compare the neutralizing antibody titers induced by CoronaVac and convalescent COVID-19 patients in parallel, however, we conducted this detection of convalescent serum specimens with same procedure performed in this phase 2 trial.

In conclusion, favorable safety and immunogenicity of CoronaVac was demonstrated on both schedules and both dosages in this phase 2 clinical trial, which support the conduction of phase 3 trial with optimum schedule/dosage per different scenarios. Currently, our first priority is to evaluate the protective efficacy of the 3 μg dosage under Day 0,14 schedule. Moreover, Day 0,28 schedule with 3 μg vaccine will also be adopted in our future phase 3 clinical trials.

## Data Availability

All the data in this manuscript is available.

## Clinical Trial number

NCT04352608

## Funding Project

National Key Research and Development Program (2020YFC0849600)

Beijing Science and Technology Program (Z201100005420023)

